# Reproducibility and reporting practices in COVID-19 preprint manuscripts

**DOI:** 10.1101/2020.03.24.20042796

**Authors:** Josh Sumner, Leah Haynes, Sarah Nathan, Cynthia Hudson-Vitale, Leslie D. McIntosh

## Abstract

The novel coronavirus, COVID-19, has sparked an outflow of scientific research seeking to understand the virus, its spread, and best practices in prevention and treatment. If this international research effort is going to be as swift and effective as possible, it will need to rely on a principle of open science. When researchers share data, code, and software and generally make their work as transparent as possible, it allows other researchers to verify and expand upon their work. Furthermore, it allows public officials to make informed decisions. In this study, we analyzed 535 preprint articles related to COVID-19 for eight transparency criteria and recorded study location and funding information. We found that individual researchers have lined up to help during this crisis, quickly tackling important public health questions, often without funding or support from outside organizations. However, most authors could improve their data sharing and scientific reporting practices. The contrast between researchers’ commitment to doing important research and their reporting practices reveals underlying weaknesses in the research community’s reporting habits, but not necessarily their science.

## Introduction

Since December 2019, more than 350,000 cases of the novel coronavirus have been reported globally, and more than 15,000 people have died.^1^ The actual number of cases is likely much higher, and the disease is still in its early phases, meaning that COVID-19 could have a staggering human toll.^2^ No matter its ultimate trajectory, the virus has already sparked an economic crisis, which will have its own implications for global justice.^3^

More now than ever, scientists and public officials need access to good, accurate information to inform their research and decision-making. Top researchers in affected nations have swiftly taken up the charge, researching the virus, its trajectory, and best practices for treating and preventing the spread of the virus. But scientists’ task does not end with analyzing results. Here at Ripeta, we have always recognized that *open* science supports *good* science, so this outbreak has heightened the importance of transparently sharing data, analysis methods, software, and code. When research is truly reproducible, it allows researchers to evaluate and expand upon each other’s work.^4^ Right now, that could mean lives saved. And, the easier it is for researchers to replicate each other’s work, the more quickly we will learn about COVID-19. We echo the sentiments of Wellcome Trust, which has urged “researchers, journals and funders to ensure that the research findings and data relevant to this outbreak are shared rapidly and openly to inform the public health response and help save lives.”^5^

The objective of this work was to classify preprints for eight different reproducibility and integrity criteria. This report provides aggregated data on 535 preprint manuscripts to explore the state of open science during this pandemic, and gives rapid initial screenings for editors and peer-reviewers to use. In providing this information, we hope to encourage best practices both now and in the future as well.

## Methods

On March 16, 2020, we harvested all preprint articles from the medRxivand bioRxiv databases related to COVID-19, for a total of 535 manuscripts. We then evaluated these articles using R statistical software version 3.5.1 for eight key reproducibility and integrity criteria: study purpose, data availability statement, data location, study location, author review, ethics statement, funding statement, and code availability (see Table 1).

**Table 1.** The Ripeta team analyzed eight reproducibility criteria, producing binary and found-value responses.

| **Transparency Criteria** | **Definition** | **Response Type** |
| --- | --- | --- |
| Study Purpose | A concise statement in the introduction of the article, often in the last paragraph, that establishes the reason the research was conducted. Also called the study objective. | Binary |
| Data Availability Statement | A statement, in an individual section offset from the main body of text, that explains how or if one can access a study's data. The title of the section may vary, but it must explicitly mention data; it is therefore distinct from a supplementary materials section. | Binary |
| Data Location | Where the article's data can be accessed, either raw or processed. | Found Value |
| Study Location | Author has stated in the methods section where the study took place or the data's country/region of origin. | Binary; Found Value |
| Author Review | The professionalism of the contact information that the author has provided in the manuscript. | Found Value |
| Ethics Statement | A statement within the manuscript indicating any ethical concerns, including the presence of sensitive data. | Binary |
| Funding Statement | A statement within the manuscript indicating whether or not the authors received funding for their research. | Binary |
| Code Availability | Authors have shared access to the most updated code that they used in their study, including code used for analysis. | Binary |

The Ripeta team analyzed the manuscripts using the Ripeta application, which leverages natural language processing (NLP) to identify and extract key pieces of text from scientific articles. Ripeta has developed several NLP models, each tuned to a specific [reproducibility criterion](https://www.ripeta.com/uploads/7/4/8/7/7487334/ripeta_approach_and_criteria_definitions.pdf). Trained to read like humans, these NLP models scan articles for seed phrases and terms that indicate the presence of their respective reproducibility criteria. Ripeta has not yet developed NLP models for all of the criteria analyzed in this study, so certain criteria required manual searches for this report. Furthermore, to ensure complete accuracy for this study, we conducted manual checks for all criteria.

For five of the criteria, the team produced binary, yes–no responses, checking for the presence but not the quality of the criteria. These criteria were study purpose, data availability statement, ethics statement, funding statement, and code availability. For study location, the team recorded both binary and text responses to indicate where the study took place. Finally, the team evaluated data location and author review and reported found values for each manuscript (See Tables 2 and 3). For author review, we created a ranking system. However, for data location, we classified data locations but did not rank them (see Table 2). Though we believe that some of these locations are easier to access and tend to have more complete data, we did not rank data locations because we recognize the need for sensitive data to be restricted for ethical reasons. However, we generally find that data stored in external repositories are more complete and are more reliably accessible. Readers can access more information about the merits and drawbacks of different data locations in the [Ripeta Approach and Criteria Definitions](https://www.ripeta.com/uploads/7/4/8/7/7487334/ripeta_approach_and_criteria_definitions.pdf) document.

**Table 2.** Data location ranking system for non-sensitive data

|  |
| --- |
| Data location not stated |
| Not publicly available |
| Data are listed as available; unable to access |
| Restricted |
| All in paper |
| All in paper or supplementary files |
| From author upon request |
| Online |
| External repository on restricted basis |
| External repository |

**Table 3.** Author review ranking system

| Tier 1 | Tier 2 | Tier 3 | Tier 4 |
| --- | --- | --- | --- |
| Author used personal email address as primary contact | Author listed but did not fill out ORCID as primary contact | Author used institutional email address as primary contact | Author listed a filled-out ORCID as primary contact |

**Table 4.**
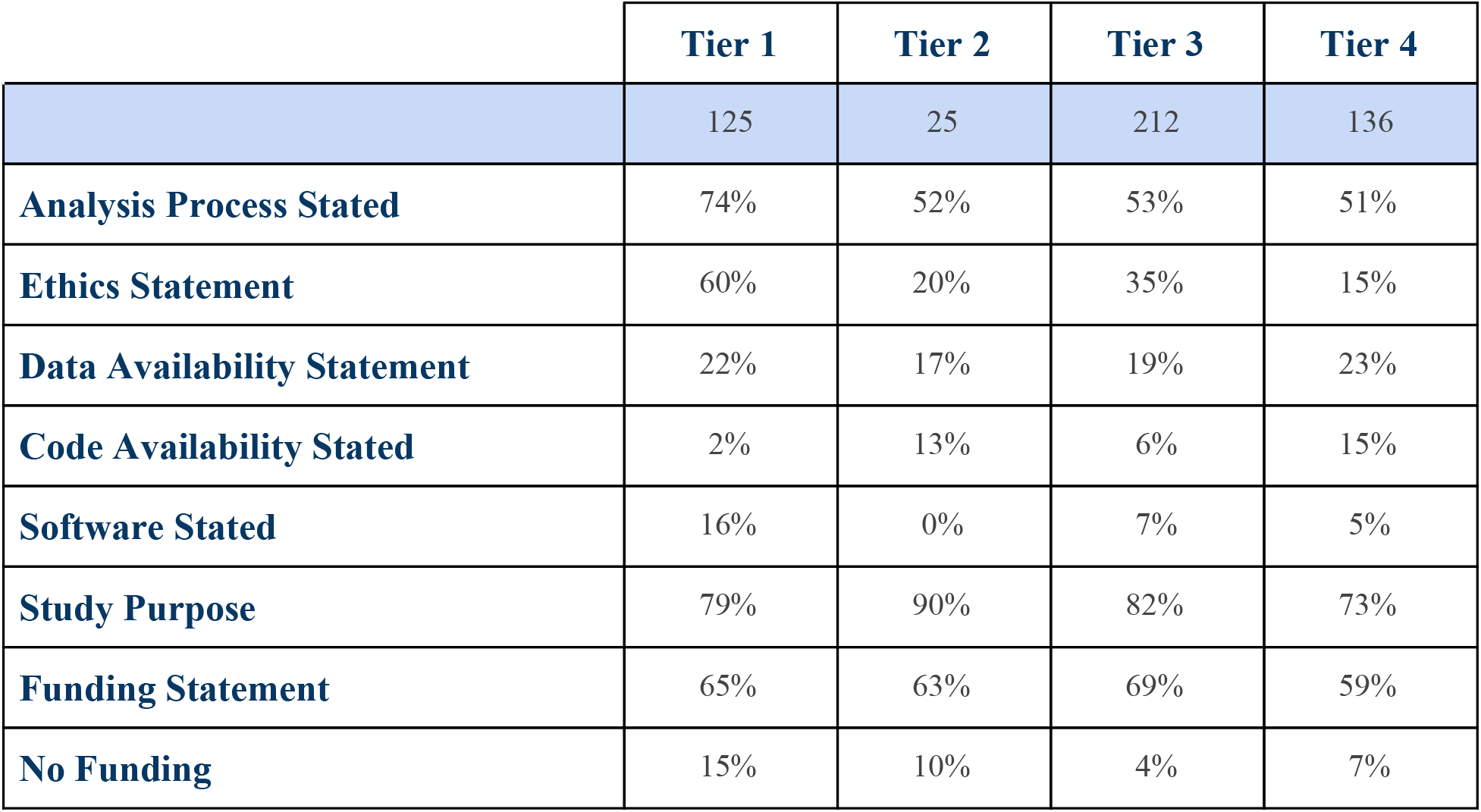
Disaggregated data for each transparency criteria by Author Tier.

Finally, the Ripeta team aggregated these data to obtain total and percent counts for each of the eight variables. Each of the variables were also separately analyzed by author tier, except for data location due to the small number of manuscripts in each possible data location.

## Results

Data for the 535 articles came from thirteen countries, with 85% of articles using data out of China (See Figure 1). Of those articles with funding statements, 8% reported that they did not receive funding for their work (See Figure 11).

**Figure 1.**
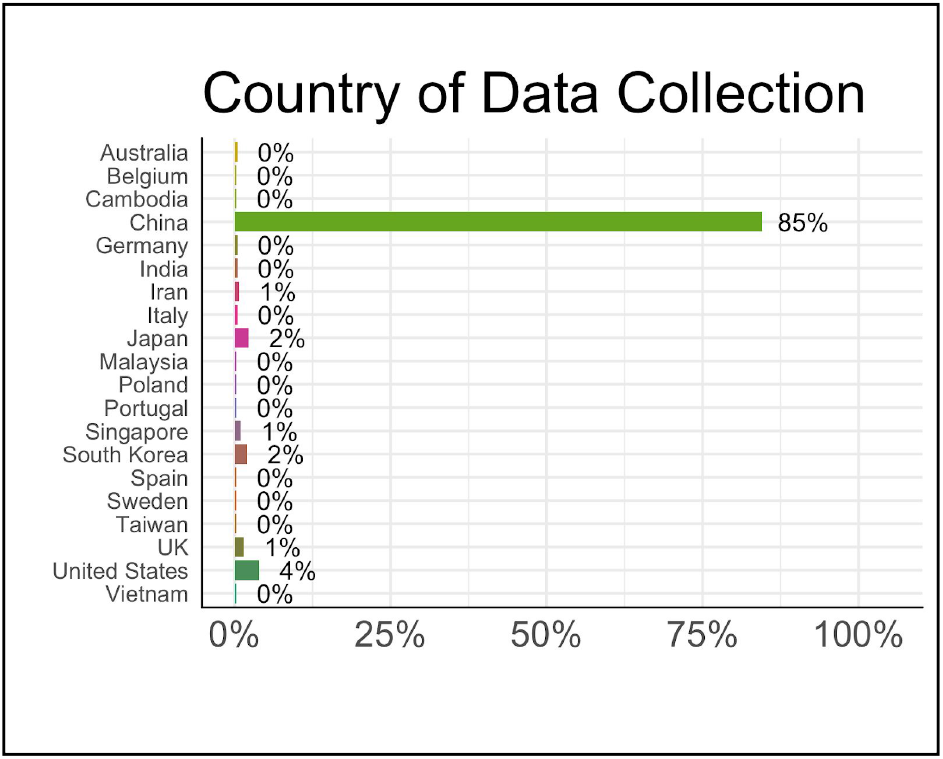
County in which data was collected, by percentage

Reporting practices varied widely by criteria. Authors most commonly included a description of their analysis processes (57%), a study purpose (79%), and a funding statement (65%). By contrast, only 8% (n=40) of articles made their code available, 21% (n=103) had data availability statements, 36% (n=172) had ethics statements, and 8%(n=41) stated what software they used. Furthermore, only 26% (n=142) of authors used a completed ORCID as their contact information, 41% (n=221) used an institutional email, 6% (n=30) listed an uncompleted ORCID, and 27% (n=138) used a personal email address. Finally, of those papers with data availability statements, only 11% (n=14) shared their data in an external repository, the preferred method of sharing data. (See Figures 2-11).

**Figure 2.**
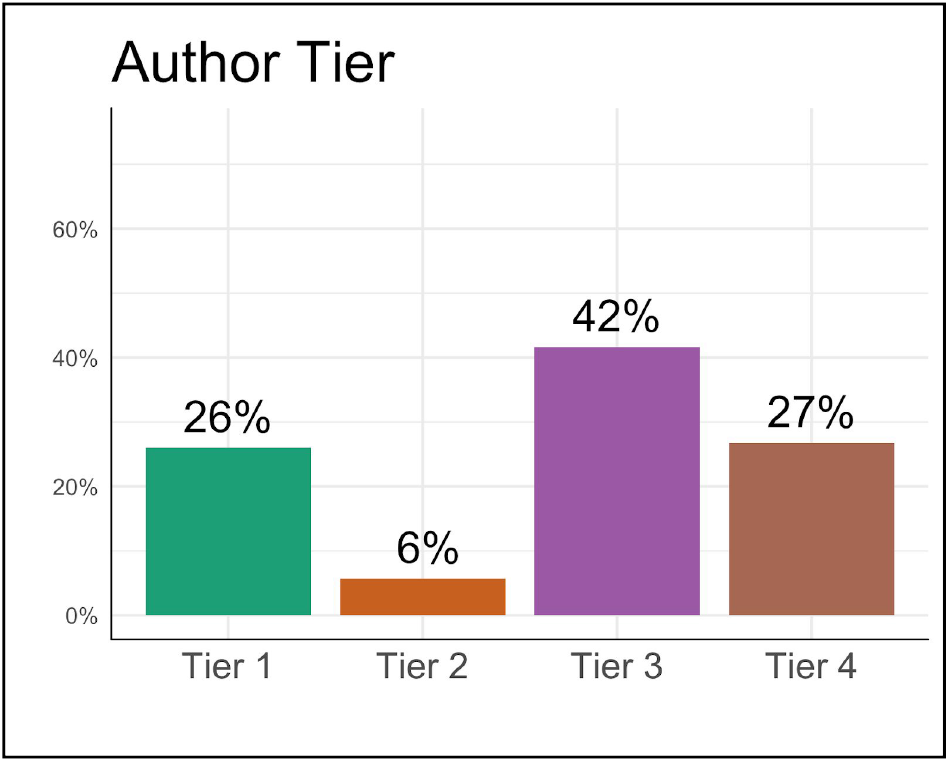
Author contact information tiers, by percentage

**Figure 3.**
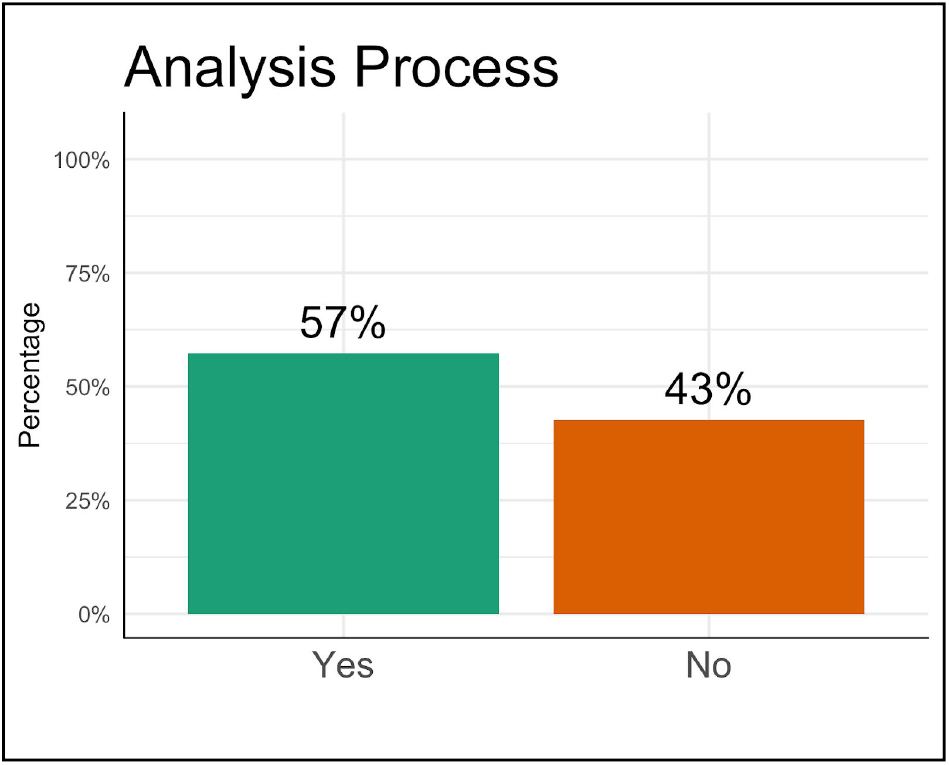
Percentage of papers that articulate an analysis process

**Figure 4.**
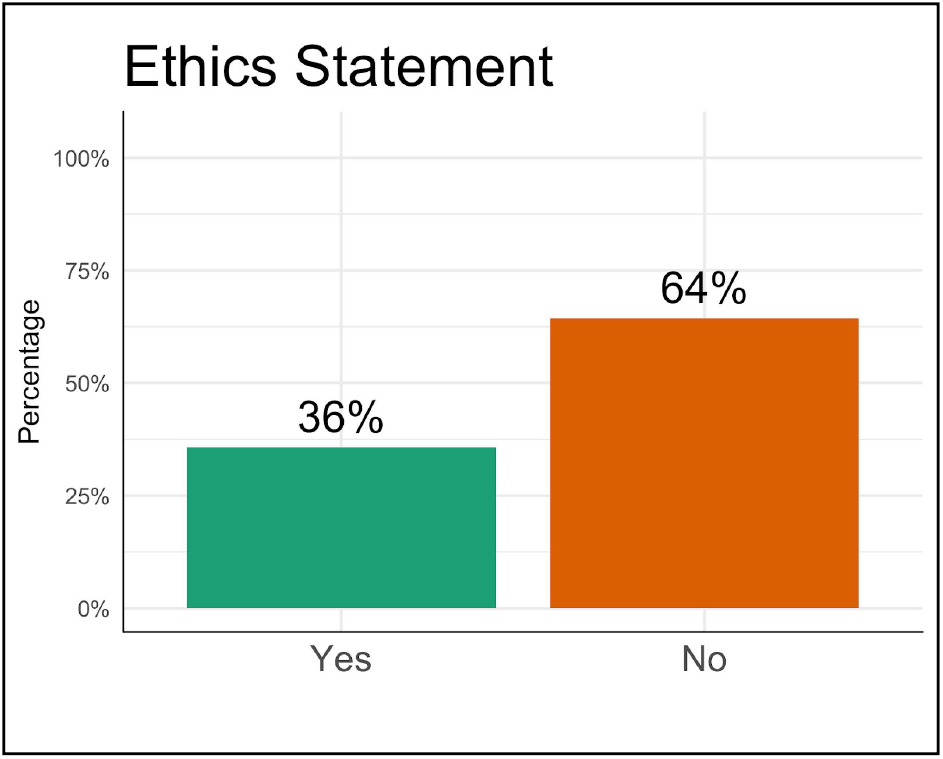
Percentage of papers with an ethics statement

**Figure 5.**
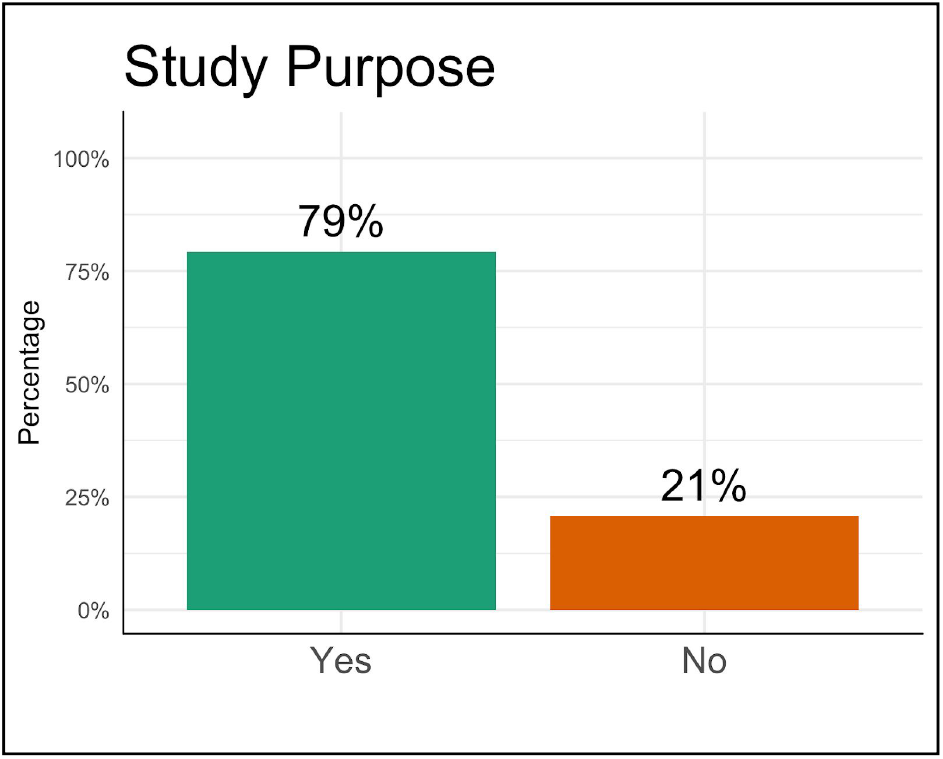
Percentage of papers articulating a study purpose, study objective, or hypothesis

**Figure 6.**
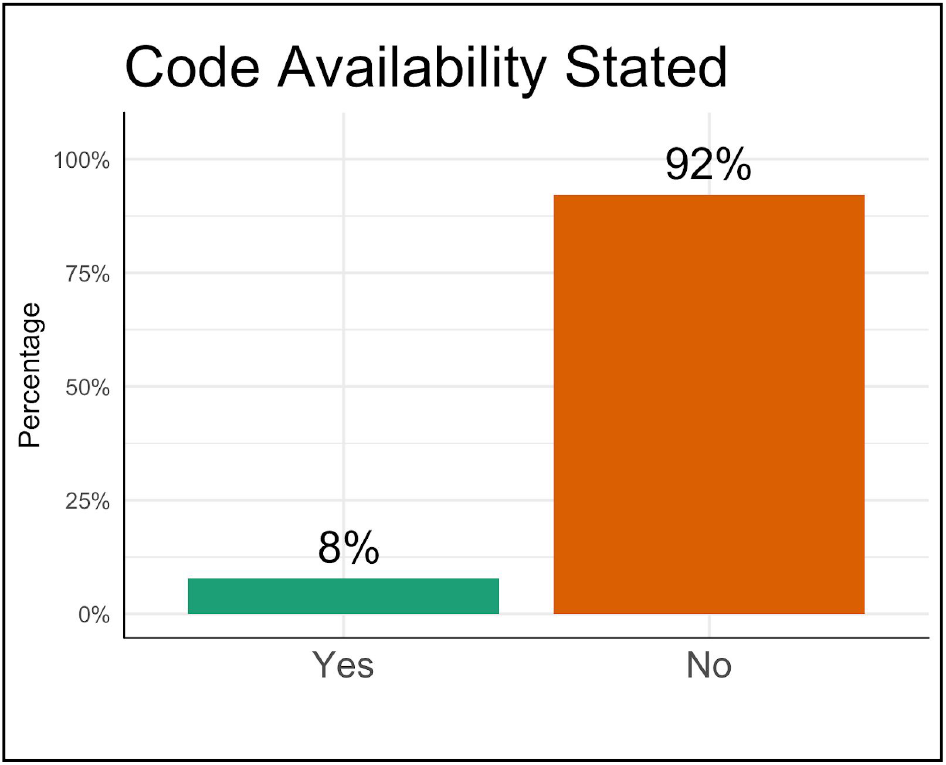
Percentage of papers that state code availability

**Figure 7.**
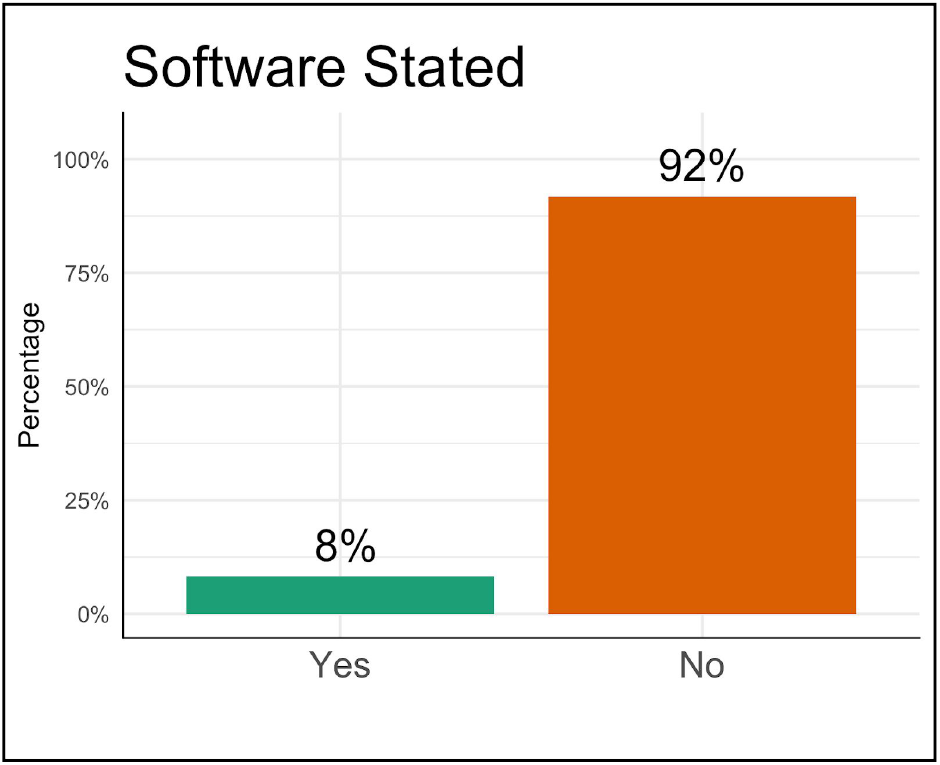
Percentage of papers that state the software they used

**Figure 8.**
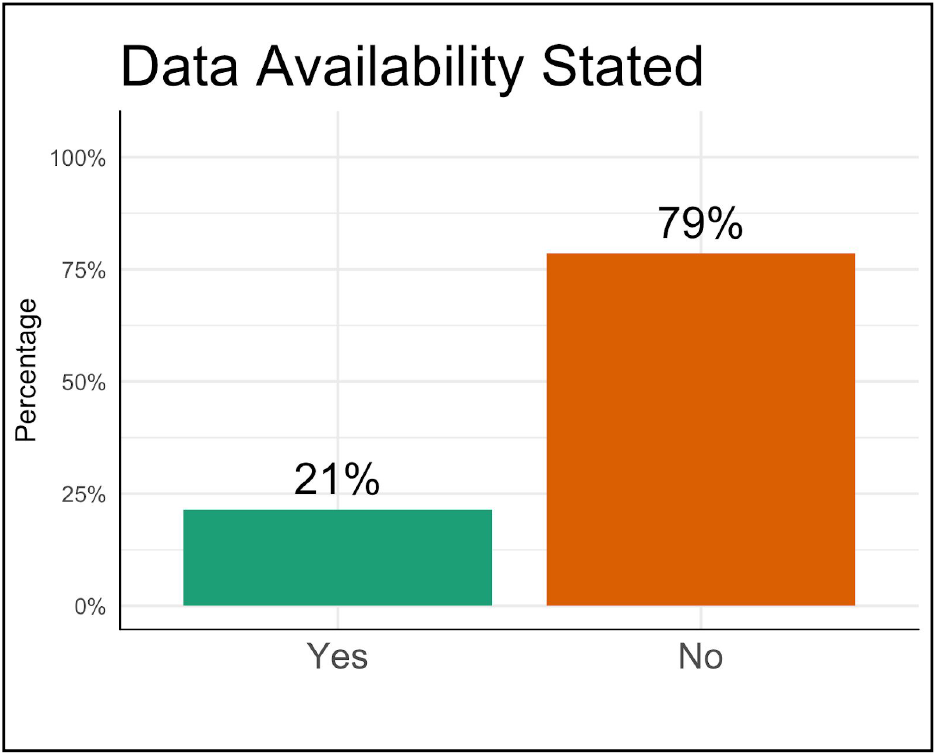
Percentage of papers with a data availability

**Figure 9.**
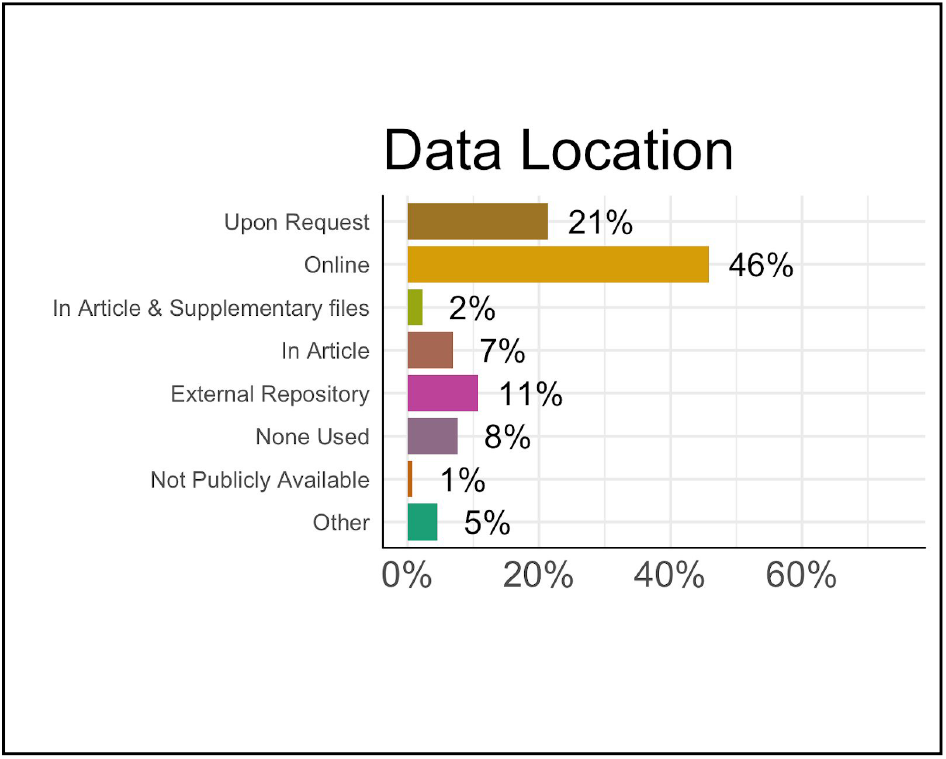
Where articles with a DAS made data available

**Figure 10.**
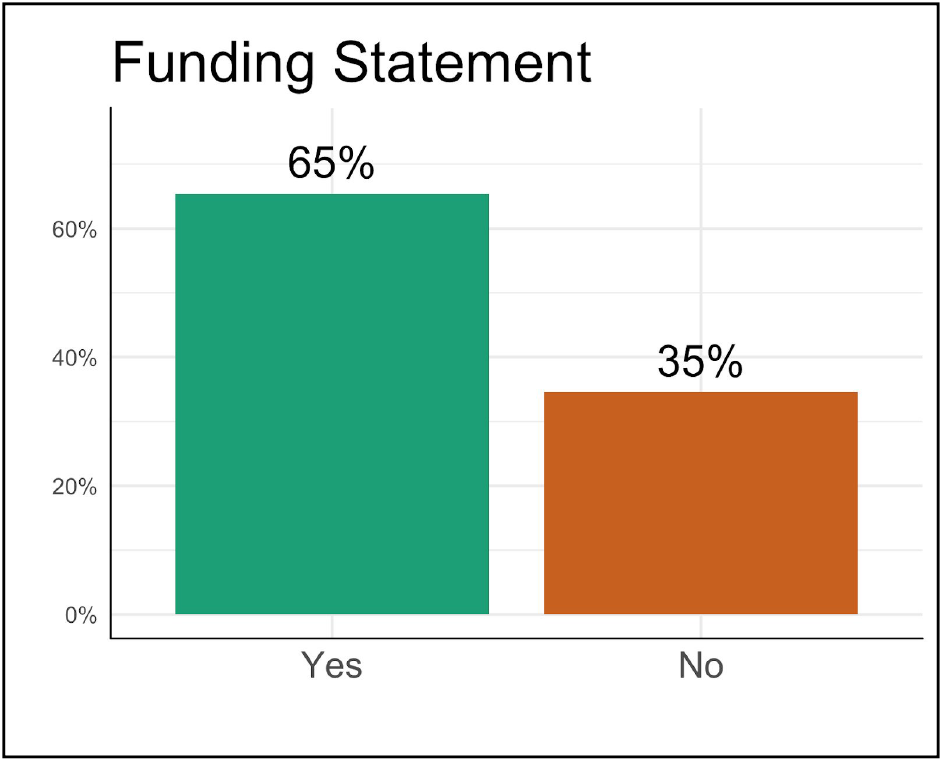
Percentage of papers with a funding statement

**Figure 11.**
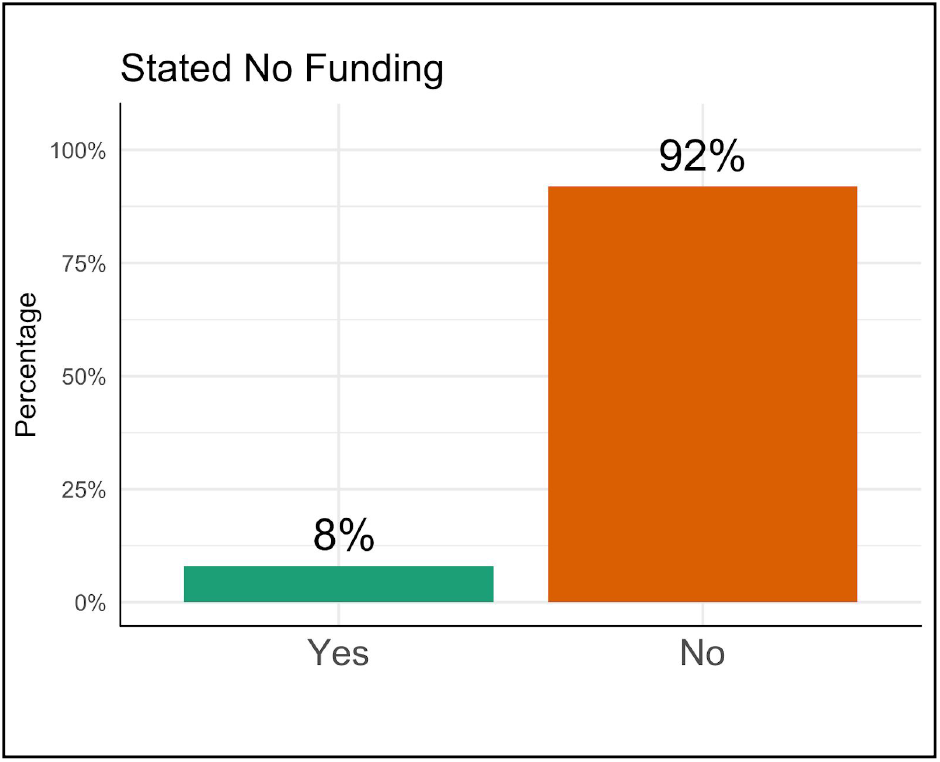
Percentage of papers with a funding statement that stated that they did not receive funding.

Though not part of the study purpose, we were curious to know if there were differences in transparency practices based on how authors identified themselves. When separated by author tier, we found significant differences for all variables except for data availability statements. Tier 1 authors, those authors providing a populated ORCID, engaged in transparency practices at a higher rate than other authors for four of the eight criteria: analysis process stated, ethics statement, software stated, and no funding. For two of the criteria, data availability statement and funding statement, there was no statistically significant difference between Tier 1 and the tier with the highest percent engagement. For the remaining two criteria, code availability stated and study purpose, Tier 1 authors complied at a significantly lower rate than the tier with the highest level of engagement. (See Table 5 for full comparisons).

Tier 1 authors reported that they had not received funding at a statistically higher rate than Tier 3 (p < 0.0006) and 4 authors (p < 0.02). Tier 1 authors also included an ethics statement at a statistically higher rate than all other authors (p < 0.0002 for each) Tier 3 authors included an ethics statement at a statistically higher rate than Tier 2 (p < 0.03) and 4 authors (p < 007). Tier 1 stated their software at a higher rate than all other authors (p < 0.007 for each) and stated their analysis processes at a higher rate than all other authors (p < 0.03 for each). Tier 2 authors stated a study purpose at a statistically higher rate than all other authors (p < 0.05 for each). Tier 3 authors included a funding statement at a significantly higher rate than Tier 4 authors (p < 0.05). Interestingly, Tier 1 authors reported their code at a significantly lower rate than all other authors (p < 0.04 for each), and Tier 4 authors additionally reported their code at a significantly higher rate than Tier 3 authors (p < 0.007).

## Discussion

The COVID-19 pandemic has laid bare the medical systems of affected nations, exposing their strengths and weaknesses. Similarly, it has revealed patterns in the scientific community. In both the medical and research communities, COVID-19 has revealed the devoted, quality work of individuals but the many challenges of the larger systems that those individuals operate within.

Since the outbreak of COVID-19 in 2019, researchers around the world have started exploring important health questions related to the virus, often without funding or outside support. Our research found that 8% of preprint articles received no funding. Furthermore, several new preprint articles related to COVID-19 are uploaded to medRxiv and bioRxiv each day. These actions reveal the dedication of scientists to provide their scientific information to decision-makers as quickly as possible. Yet, we found that most authors are not employing best reporting practices, despite the importance of open science during a health crisis. Many authors did not make their data readily accessible, did not share code, and did not engage in other important transparency practices. At a time when many scientists are going above and beyond to promote their research, these findings suggest that the scientific community may not recognize the role that certain criteria play in reproducibility and may not appreciate the extent to which open science promotes innovation and advancement. It may also signal that it is simply difficult to improve the reporting and data sharing practices within science.

In particular, the large discrepancies between criteria suggest that authors care about good reporting practices but that certain criteria are far more salient and highly valued than others. For instance, the vast majority of authors included a study purpose, yet almost no authors reported their code. Furthermore, almost all authors provided a funding statement, and over half of authors described their analysis processes. Perhaps most importantly, only 21% of authors included data availability statements, and only 11% of those made their data available in external repositories. Though we found that the data for 46% of those articles were available online, many of these studies used pre-existing online data, meaning that they did not actually make new data available. These low numbers suggest that some authors might not recognize the importance of data sharing, believing that they only need to share their methods in order to make their studies reproducible. Overall, authors who engaged in more reporting practices may have simply been exposed to more information about how to promote transparency. Conversely, authors who engaged in fewer of the practices may have been confused by conflicting advice in the myriad of available author guidelines.^6^ Thus, our findings suggest the need to make clear cases for a handful of the most important reporting practices as often and as publicly as possible.

All major publishers and funding organizations have author guidelines that stress the importance of open reporting – and some even require certain best practices – yet most published articles do not comply with these guidelines, as they are not mandated and are time-consuming to check. Though authors may care about reporting practices, they may not know which ones are most important, and they may choose to focus their energy on the content of their research. Furthermore, the peer review process is labor intensive and time consuming, and reviewers may understandably focus on process, results, and content more than specific reporting guidelines. The scientific community can combat this issue by implementing automated reviews when possible. As shown in this study, there are many checks for certain reproducibility and integrity criteria that have been and can be automated. It took approximately 30 minutes to do automated checks for all 535 papers, while our manual checks took approximately 5 minutes per paper, culminating in dozens of hours of work. Thus, automation would make reporting checks feasible by drastically reducing the time it takes to conduct them, allowing expert reviewers to focus on evaluating the science itself. Before proceeding to peer review, authors would need to address any transparency concerns and conduct a second automated check.

Whichever interventions the scientific community eventually implements, we hope that those researchers committed to combating the COVID-19 outbreak will recognize the inherent importance of open science. Many of these researchers seem to be motivated by altruism rather than career, so we must continue to highlight those reporting practices that will facilitate reproducibility and falsifiability. Finally, we must reiterate the general importance of open science: that our best work comes from the knowledge we create together, across borders and with collaborators we may never know exist.

## Data Availability

Data and code are available on figshare.
Data: 10.6084/m9.figshare.12026364
Code: 10.6084/m9.figshare.12026370

https://figshare.com/articles/Covid_Preprint_Data/12026364

https://figshare.com/articles/Covid_Preprint_-_R_Code/12026370

## Data and Code Availability Statement

Data and code are available on figshare.

Data: 10.6084/m9.figshare.12026364

Code: 10.6084/m9.figshare.12026370

## Ethics Statement

No ethics approval was needed for this study.

## Author Contributions

JS Data analyses; manual annotations

LH Completed manual annotations

SN Manuscript drafting and editing

CHV Study design and manuscript editing

LDM Study design; analyses oversight; manuscript editing

## Footnotes

1 Coronavirus disease (COVID-19) outbreak. (2020, March 20). *World Health Organization*. Retrieved from https://www.who.int/emergencies/diseases/novel-coronavirus-2019

2 Liu, Y., Gayle, A A., Wilder-Smith, A., Rocklöv, J. (2020). The reproductive number of COVID-19 is higher compared to SARS coronavirus. *Journal of Travel Medicine, 27(2)*. Retrieved from https://doi.org/10.1093/jtm/taaa021

3 McKibbin, W. J. & Fernando, R. (2020). The Global Macroeconomic Impacts of COVID-19: Seven Scenarios. CAMA Working Paper No. 19/2020. Retrieved from http://dx.doi.org/10.2139/ssrn.3547729

4 Begley, C. G., & Ioannidis, J. P. A. (2015). Reproducibility in science: Improving the standard for basic and preclinical research. *Circulation Research, 116*(1), 116-126. doi:10.1161/CIRCRESAHA.114.303819

5 Sharing research data and findings relevant to the novel coronavirus (COVID-19) outbreak. (2020, January 30). *Wellcome Trust*. Retrieved from https://wellcome.ac.uk/press-release/sharing-research-data-and-findings-relevant-novel-coronavirus-covid-19-outbreak

6 Malički M, Aalbersberg IJ, Bouter L, ter Riet G (2019) Journals’ instructions to authors: A cross-sectional study across scientific disciplines. PLoS ONE 14(9): e0222157. https://doi.org/10.1371/journal.pone.0222157

## References

Begley, C. G., & Ioannidis, J. P. A. (2015). Reproducibility in science: Improving the standard for basic and preclinical research. Circulation Research, 116(1), 116–126. doi:10.1161/CIRCRESAHA.114.303819

Coronavirus disease (COVID-19) outbreak. (2020, March 20). World Health Organization. Retrieved from https://www.who.int/emergencies/diseases/novel-coronavirus-2019

Liu, Y., Gayle, A A., Wilder-Smith, A., Rocklöv, J. (2020). The reproductive number of COVID-19 is higher compared to SARS coronavirus. Journal of Travel Medicine, 27(2). Retrieved from https://doi.org/10.1093/jtm/taaa021

Malički M, Aalbersberg IJ, Bouter L, ter Riet G (2019) Journals’ instructions to authors: A cross-sectional study across scientific disciplines. PLoS ONE 14(9): e0222157. https://doi.org/10.1371/journal.pone.0222157

McKibbin, W. J. & Fernando, R. (2020). The Global Macroeconomic Impacts of COVID-19: Seven Scenarios. CAMA Working Paper No. 19/2020. Retrieved from http://dx.doi.org/10.2139/ssrn.3547729

McIntosh LD, Juehne A, Vitale CRH, et al. Repeat: a framework to assess empirical reproducibility in biomedical research. BMC Medical Research Methodology. 2017 Sep;17(1):143. DOI: 10.1186/s12874-017-0377-6.

Sharing research data and findings relevant to the novel coronavirus (COVID-19) outbreak. (2020, January 30). Wellcome Trust. Retrieved from https://wellcome.ac.uk/press-release/sharing-research-data-and-findings-relevant-novel-coronavirus-covid-19-outbreak

